# Mild cognitive impairment is associated with skeletal muscle mitochondrial deficits

**DOI:** 10.1101/2020.07.20.20158022

**Authors:** Jill K. Morris, Colin S. McCoin, Kelly N. Fuller, Casey S. John, Heather M. Wilkins, Xiaowan Wang, Palash Sharma, Jeffrey M. Burns, Eric D. Vidoni, Jonathan D. Mahnken, Russell H. Swerdlow, John P. Thyfault

## Abstract

Alzheimer’s Disease (AD) is associated with insulin resistance and low cardiorespiratory fitness, suggestive of impaired skeletal muscle mitochondrial function. We examined if individuals with Mild Cognitive Impairment (MCI), the earliest phase of AD-related cognitive decline, exhibit reduced skeletal muscle mitochondrial function, and if AD medication impacted outcomes. We present data from 50 individuals, including cognitively healthy older adults (CH; n=24) 60+ years of age and clinically diagnosed MCI subjects (n=26). MCI subjects were sub-divided into two groups; no AD medication (MCI; n=11), or AD medication treated (MCI+med; n=15). A skeletal muscle biopsy (vastus lateralis) was obtained and mitochondrial respiratory kinetics was measured in permeabilized muscle fibers. MCI subjects exhibited lower lipid-stimulated skeletal muscle mitochondrial respiration (State 3, ADP-stimulated) than both CH individuals (p=0.043) and medication-treated MCI subjects (p=0.006). MCI also exhibited poorer mitochondrial coupling control compared to CH subjects (p=0.014), while MCI+med and CH subjects did not differ. Compared to CH individuals, skeletal muscle mitochondrial leak control ratio was lower for the MCI+med group (p=0.008) and trended lower for non-medicated MCI (p=0.06), which suggests greater mitochondrial uncoupling in MCI. Skeletal muscle mitochondrial respiration is impaired in untreated MCI but normalized in medication-treated MCI participants while mitochondrial leak control is impaired regardless of medication status. These results provide further evidence that systemic mitochondrial deficits occur in the very early stages of AD, and that mitochondrial function is partially influenced by AD medication. Further analysis for a role of muscle mitochondria in the progression of early AD is warranted.

## Introduction

Emerging evidence suggests that changes in energy metabolism may play a role in the development and progression of Alzheimer’s Disease (AD). Results show that mitochondrial function is compromised systemically in AD, in tissues ranging from skeletal muscle to brain,^1^and we have shown that AD subjects have both reduced aerobic capacity ^2^ and reduced insulin sensitivity^3^ compared to cognitively healthy older adults. There is also evidence that mitochondrial dysfunction in skeletal muscle is a cause of whole body insulin resistance,^4–6^ a recognized risk factor for AD. ^7–20^ Moreover, skeletal muscle mitochondrial content and functional capacity plays a key role in maximal exercise capacity and cardiorespiratory fitness. However, skeletal muscle mitochondrial function has never been directly assessed in cognitively impaired individuals. Given that skeletal muscle tissue comprises 30-40% of body mass and declines with aging,^21^ metabolic deficits in this tissue have a profound impact on systemic metabolism and the development of age-related chronic disease states.

The overall goal of this study was to test the hypothesis that individuals in the earliest stage of AD-related cognitive decline, Mild Cognitive Impairment (MCI), possess reduced mitochondrial function and specifically, reduced State 3 (ADP-stimulated) skeletal muscle mitochondrial respiratory capacity compared to cognitively healthy elderly adults (CH). We also postulated that differences in respiratory capacity could be driven by AD genetic risk (Apolipoprotein ε4; APOE4)) or non-genetic risk (overweight/obesity or physical activity). However, emerging literature also suggests that AD medications influence mitochondrial function. Because over half (58%) of MCI subjects in our study were actively using AD medication, we also characterized skeletal muscle mitochondrial effects in the presence and absence of medication treatment. Our results show a clear effect of MCI status being linked to compromised mitochondrial function in skeletal muscle that is partially modified by medication status.

## Methods

### Ethical approval and recruitment

This study was approved by the University of Kansas Medical Center’s Institutional Review Board (IRB # 140787). All participants in this study provided informed consent according to institutional guidelines and in accordance with the Declaration of Helsinki. Participants were recruited by the KU Alzheimer’s Disease Center (KU ADC) recruitment division as previously described.^57^ 180 qualifying individuals were pre-screened for this study by the KU ADC recruitment operations team. 58 individuals were selected for additional in-depth screening by the study coordinator, with 52 participants enrolled in the study. One individual was withdrawn from the study prior to the muscle biopsy due to safety labs out of range and one individual was excluded from analysis due to the muscle sample being compromised prior to analysis. We thus report results from 50 participants.

All individuals were over 60 years of age and on stable medication doses for at least 30 days. Diagnostic inclusion criteria included no prior memory complaints (CH older adults), or MCI diagnosed by a clinician and verified with medical records. Individuals were excluded from participating if they had moderate or severe AD, other neurological disorders that could impair cognition, evidence of bleeding disorders during screening (elevated PT/PTT values), prior bleeding problems or use of anti-platelet medication, Warfarin, or any other anticoagulant, clinically significant disease, clinically significant psychiatric disorder, systemic illness or infection likely to affect safety, clinically-evident stroke, myocardial infarction or coronary artery disease in the last 2 years, insulin dependent diabetes, or significant pain or musculoskeletal symptoms that would affect safety. One individual in the CH group had a diagnosis of Type 2 Diabetes but was not using diabetic medication.

Interested participants completed a brief phone screen to verify safety to complete visits. Participants that appeared eligible were scheduled an in-person visit. A screening blood panel was also performed prior to the muscle biopsy to verify that no blooding disorders were present. Individuals completed 2 study visits: Visit 1 (Graded exercise test; GXT, and dual energy x-ray absorptiometry (DXA) scan), and Visit 2 (fasting blood draw, muscle biopsy).

### Medications

Medication use was verified by medical record. All individuals in the study were required to be on stable medication doses for greater than 30 days prior to enrollment. Fifteen MCI subjects (58% of study subjects with MCI) were routinely taking medication for mild memory complaints. All 15 of these individuals were using the cholinesterase inhibitor donepezil (10mg/day; 12 or 5mg/day; 3, due to inability to tolerate a 10mg/day dose). Of these 15 subjects, 5 individuals were also taking memantine, an NMDA receptor antagonist (5- 28mg/day).

### Anthropometric measures

Individuals reported to the KU Clinical and Translational Research Unit (CTSU) for Visit 1 following an overnight fast. After a 5 minute rest period, vital signs were obtained. We measured height in cm and total body mass using a digital scale accurate to 0.1kg (Seca Platform Scale, model 707). Waist and hip circumference were also measured. Subjects were asked to void and evaluated using a DXA scan (Lunar Prodigy, version 11.2068) to determine lean mass, fat mass, and bone mineral density.

### Cardiorespiratory fitness testing

A GXT was performed according to a modified Bruce protocol on a treadmill as previously described to determine cardiorespiratory fitness.^58^ Subjects were attached to a 12- lead electrocardiograph to monitor cardiac rhythm, and a non-rebreathing mask to continuously capture oxygen and carbon dioxide for 3 minutes while seated and relaxed at rest. This allowed for calculation of a resting Respiratory Exchange Ratio (RER). To begin the test, each participant mounted a treadmill and began walking with speed and incline gradually increasing with each 2 minute stage as previously reported.^59^ Blood pressure and rating of perceived exertion were collected at the end of each stage. The test was terminated if the participant reached volitional fatigue or met the absolute test termination criteria (RER>=1.1, RPE 17, plateau in VO_2_/100ml change, 90% HRmax). Maximal oxygen uptake during the GXT was calculated relative to whole body mass (mL/kg/min). Our group has previously validated VO_2_ peak testing in both AD subjects and in CH older adults.^58,60,61^

### Accelerometry

At Visit 1, individuals were given a wrist-worn Actigraph GT9X watch (Sample Rate 30Hz) and asked to wear the watch for 7 days to capture both weekday and weekend physical activity and sedentary behavior patterns. Accelerometers were returned to the study team during Visit 2 for data analysis. Wear validation was performed using a computer algorithm ^62^ with a minimum of 600 mins wear time per day and 4 valid days required for inclusion of data. No individuals were excluded from analysis due to inadequate wear time. 1 subject was excluded from the analyses due to corrupt data. Analyses were performed using Actigraph ActiLife 6 software (v6.13.3).

### Blood biomarkers

Approximately 2 weeks following Visit 1, subjects returned to the KU Clinical Translational Science Unit (CTSU) for visit 2 following an overnight fast. Blood samples were analyzed using a lipid panel (Quest Diagnostics) to determine cholesterol HDL, LDL, LDL/HDL, and triglyceride values. Blood was collected into serum separator tubes and processed to generate serum, which was frozen at −80°C until further analysis. Serum glucose was measured using a colormetric assay (Sigma-Aldrich) and insulin was measured using ELISA (Alpco Diagnostics).

### Muscle biopsy and mitochondrial respirometry analyses

Following the visit 2 blood draw, a skeletal muscle biopsy of the vastus lateralis (~120 mg tissue) was performed. Freshly obtained muscle tissue was immediately dissected free of connective tissue and separated into segments, with a portion (~30mg) placed into ice cold buffer X (50 mM K-MES, 7.23 mM K_2_EGTA, 2.77 mM CaK_2_EGTA, 20 mM Imidazole, 20 mM Taurine, 5.7 mM Na-ATP, 14.3 mM Na-PCr, 6.56 mM MgCl_2_-6H_2_O; pH 7.1) for transport to the laboratory and analysis of respiratory kinetics.

We measured basal and ADP-stimulated respiratory kinetics utilizing lipid substrates in duplicate on permeabilized fiber bundles in an Oroboros Oxygraph-2k system (Innsbruck, Austria). All mitochondrial analyses were completed in real time within 2-3 hours of the biopsy. Multiple small fiber bundles were teased from the ~30mg muscle sample on ice using fine forceps under a dissecting scope in ice cold buffer X. Following fiber bundle preparation, bundles were placed into buffer X containing 30ug/mL saponin for 30 minutes on a rotator at 4°C, prior to washing with ice cold Buffer Z (105 mM K-MES, 30 mM KCl, 10 mM K_2_HPO_4_, 5 mM MgCl_2_-6H_2_O and 0.5% w/w fatty acid-free BSA; pH 7.1) + 0.5 M EGTA.

We used high resolution respirometry to assess O_2_ consumption rate within prepared muscle fiber bundles. Analyses were performed at 37°C in Buffer Z with 0.5 M EGTA and 20 mM creatine monohydrate. Basal respiratory kinetics were obtained in the presence of 0.01 mM blebbistatin, 0.02 mM palmitoyl CoA, 0.5 mM malate, 5 mM carnitine, and 0.018 mM palmitoylcarnitine. O_2_ was added to the chamber for an initial concentration of ~260 μM. After reaching steady state, this was followed by sequential addition of 4 mM ADP to assess State 3 respiration through Complex I, and 10 mM succinate (Complex II substrate; State 3S). We assessed uncoupled respiration by adding carbonylcyanide-p-trifluoromethoxyphenylhydrazone (FCCP, a protonophoric uncoupler) until we observed maximal respiration. Following analyses, fibers were freeze dried overnight in a lyophilizer (FreeZone 2.5L, Labconco, Kansas City, MO) and weighed using a microbalance (MX5, Mettler Toledo, Columbus, OH). Mitochondrial respiration values were normalized to dry muscle weight and are expressed as pmol·s^−1^·mg^−1^ dry weight. We calculated coupling control ratio by dividing leak respiration by phosphorylating State 3 respiration (ADP condition; L/P). We calculated leak control ratio by dividing leak respiration by FCCP stimulated respiration (L/E).

### Protein expression analyses

Frozen muscle was powdered and weighed prior to processing with a TissueLyser II bead homogenizer (Qiagen, Germantown, MD) in buffer containing 50 mM HEPES, 12 mM sodium pyrophosphate, 100 mM NaF, 10 mM EDTA, 400 μL each phosphatase inhibitor cocktail 2 and 3 (Sigma-Aldrich, St. Louis, MO), and 1% Triton X-100. Protein concentration assessed using a Pierce BCA assay (Thermo Scientific, Rockford, IL) and samples prepared for western blotting prior to separation using SDS-PAGE. Proteins were transferred to PVDF membrane prior incubation with Total OXPHOS primary antibody (Abcam, Cambridge, MA). Densitometry was measured using ImageLab 5.2.1 (BioRad, Hercules, CA) and protein loading corrected using 0.1% amido-black (Sigma-Aldrich).

### Statistical Analyses

Group differences for continuous variables were assessed using ordinary least squares regression to perform ANOVA-like analyses with adjustment for other factors. Significant findings were further explored using LSD post-hoc analyses. For an overall assessment of mitochondrial response under multiple substrate conditions, we used linear mixed modeling to account for repeated measures. Model assessment included residual analysis. Linear mixed model results assuming the normal distribution were violated with respect to the constant variance assumption. Log-transformation did not correct this problem with the residuals. We thus adjusted to use a generalized linear mixed model assuming an underlying Poisson distribution as this allowed for the variance to increase as a function of the mean, and this resolved the poor diagnostic indications from the linear mixed models. The relationship between CRF and our primary mitochondrial outcome measure of State 3 (ADP) flux was explored using linear regression. All analyses were adjusted for age, sex, APOE4 carrier status, and overweight/obesity status. Values are prented in tables as mean [SD]. Results were considered significant at *p<0.05.

## Results

### Subject Characteristics

CH, MCI, and MCI+med groups did not significantly differ by age, sex, genotype (APOE4 carrier status) or overweight/obesity status (**Table 1**). Within the MCI+med group, all participants (n=15, 100%) were using the AD medication donepezil, an acetylcholinesterase inhibitor. Of those subjects, 33% (n=5) were also using the AD medication memantine. There was no difference in years of education between groups. Scores for the Mini-Mental State Examination (MMSE) were available for all MCI subjects. MMSE scores were not different between MCI and MCI+med groups (mean MMSE score 26 for each group), and these scores are within the range commonly observed in MCI populations.^22^

**Table 1.** Subject characteristics.

Values are given as means [SD]. p<0.05 <sup>#</sup>CH vs MCI, <sup>+</sup>CH vs MCI+med, <sup>\*</sup>MCI vs MCI+med. APOE; Apolipoprotein E, BMI; body mass index, GXT; graded exercise test, RER; respiratory exchange ratio, HR; heart rate.
| Outcome | CH (n=24) | MCI (n=11) | MCI + Med (n=15) | p-value |
| --- | --- | --- | --- | --- |
| Age (y) | 72.5 [8.5] | 69.6 [7.4] | 74.9 [7.3] | 0.253 |
| Sex (#, % female) | 12 [48%] | 9 [82%] | 7 (47%) | 0.128 |
| APOE4 (#, % carrier) | 9 [36%] | 8 [73%] | 7 [47%] | 0.126 |
| Education (y) | 16.9 [1.7] | 16.9 [2.3] | 15.8 [2.3] | 0.250 |
| Overweight/Obesity (#,%) | 15 [62%] | 3 [27%] | 9 [60%] | 0.130 |
| Activity per day (counts) | 12689.8 [3470] | 14936 [3242] | 10121.0 [3014] | **0.010<sup>\*,+</sup>** |
| VO<sub>2 peak</sub> (mL/kg/min) | 26.5 [6.9] | 25.8 [4.2] | 22.8 [6.7] | 0.155 |
| GXT duration (min) | 13.1 [2.6] | 12.4 [2.5] | 9.51 [3.3] | **<0.001<sup>\*,+</sup>** |
| Resting RER | 0.831 [0.08] | 0.773 [0.04] | 0.780 [0.08] | **0.031<sup>#,+</sup>** |
| Peak HR | 159.8 [20.1] | 154.9 [15.0] | 148.7 [27.4] | 0.437 |
| BMD | 1.25 [0.19] | 1.11 [0.16] | 1.20 [0.13] | 0.228 |
| Waist Hip Ratio | 0.883 [0.12] | 0.814 [0.08] | 0.917 [0.11] | 0.276 |
| Lean mass (kg) | 47.5 [10.7] | 37.9 [10.6] | 48.3 [10.4] | 0.100 |
| Fat mass (kg) | 27.0 [9.3] | 19.5 [5.1] | 29.9 [11.2] | **0.027<sup>+</sup>** |

### Mitochondrial respiration

To test mitochondrial function, we measured respiratory kinetics in permeabilized skeletal muscle fiber bundles. Analysis of our primary outcome measure, the respiratory response to ADP, was different between groups. We observed differences in ADP-stimulated (State 3) respiratory kinetics (p=0.020; **Figure 1A**), with MCI subjects exhibiting lower mitochondrial respiration than CH (p=0.043) and MCI+Med (p=0.006) individuals. We also performed multiple additional titrations of substrates, including Succinate (State 3S) and FCCP (uncoupled respiration) to assess respiratory control under a sequence of conditions. In our analysis accounting for these repeated measures across all sequence conditions, we also found the relationship between groups differed across the sequence of conditions (p<0.001). Within condition difference between groups were detected in this repeated measures analysis for the ADP-stimulated (State 3) condition (p=0.005), and for Basal, Succinate (State 3S), and FCCP group comparison tests resulted in p=0.067, p=0.254, and p=0.073, respectively (**Figure 1B**). In this repeated measures analysis, within the ADP-stimulated condition MCI subjects again indicated lower mitochondrial respiration than CH (p=0.016) and MCI+Med (p=0.001) individuals.

**Figure 1.**
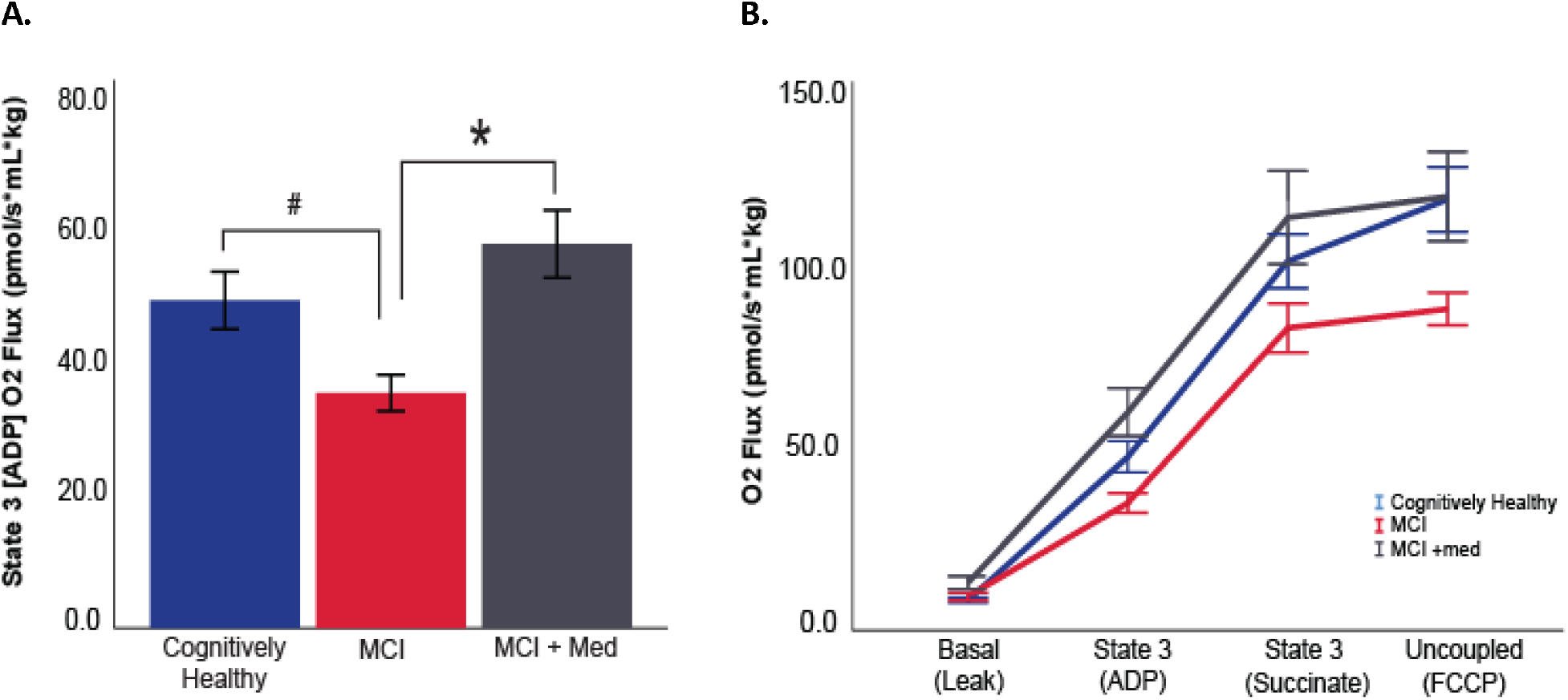
Skeletal muscle mitochondrial respiration differs between groups. Lipid (palmitoylcarnitine) stimulated skeletal muscle mitochondrial respiration was lower in MCI subjects during State 3 (ADP) compared to both other groups (A). Responses were also assessed under a sequence of conditions, and the overall group response differed across the states (p<0.001). Ordinary least squares regression was used for State 3 univariate analyses. The repeated measures overall responses to substrate conditions was assessed using linear mixed modeling. Both sets of analyses adjusted for covariates. #p<0.05 MCI no drug vs. CH. *p<0.05 MCI vs. MCI+med. CH n=24, MCI n=11, MCI+med n=15 subjects per group.

Coupling control ratio provides information on the coupling of electron flux through the electron transport chain to ATP production (lower values equal higher coupling). Coupling control ratio was significantly different between groups (p=0.045; **Figure 2A**). MCI subjects exhibited poorer coupling control compared to CH subjects (p=0.014), while MCI+med and CH subjects did not differ. Similarly, leak control ratio, which can be used to assess uncoupling during constant electron transport flow was also different between groups (p=0.019; **Figure 2B**). Leak control ratio was impaired in the MCI+med group compared to CH subjects (p=0.008) with a trend for a difference in MCI vs. CH (p=0.06), suggesting greater uncoupling of the system in MCI regardless of medication status.

**Figure 2.**
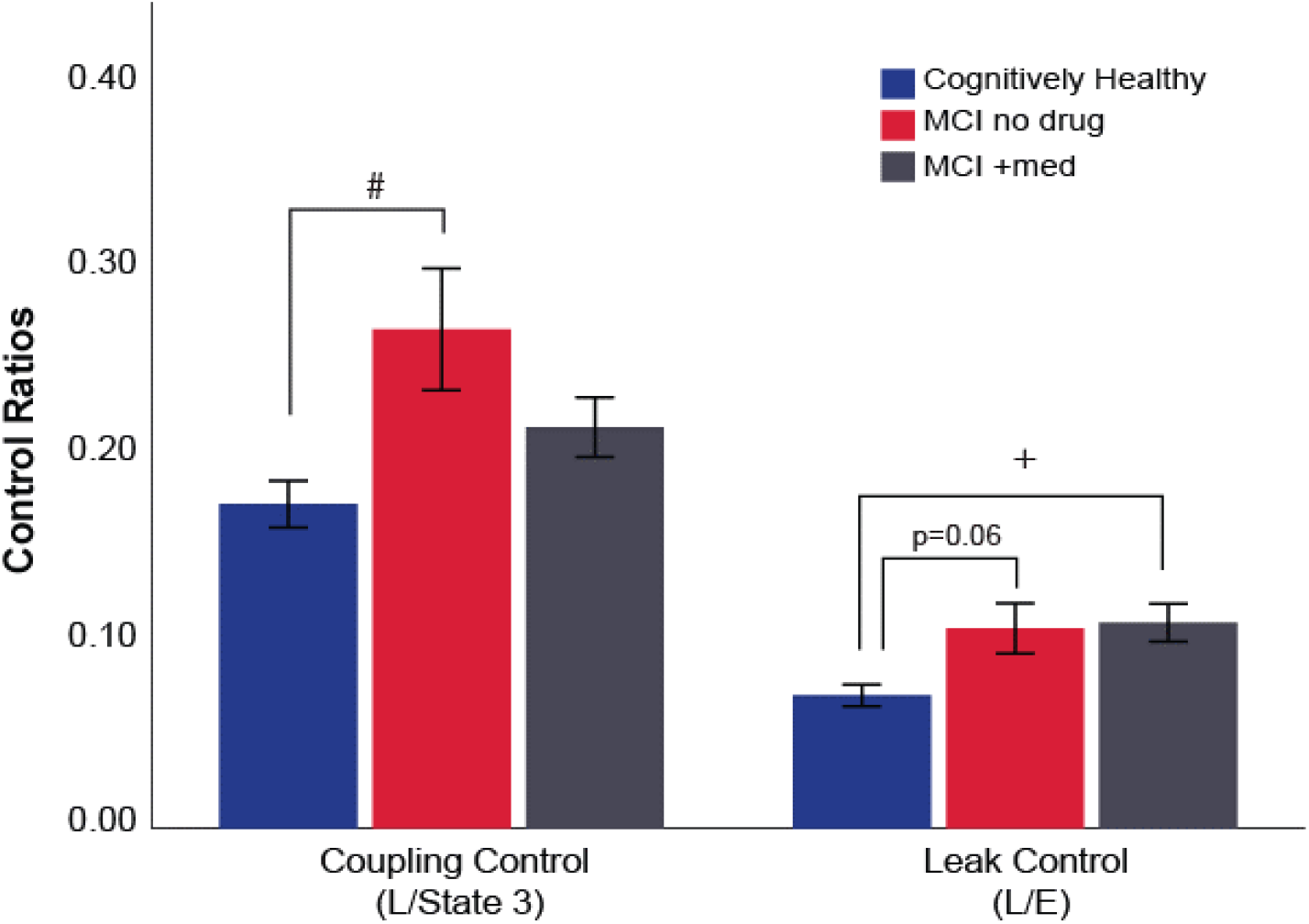
Diagnostic differences in mitochondrial control ratios. MCI subjects not treated with mediation exhibited poorer coupling control (higher control ratio) compared to CH subjects. Both MCI groups exhibited poorer leak control compared to CH individuals. Group differences were assessed using ANCOVA, adjusting for covariates. #p<0.05 CH vs MCI, +p<0.05 CH vs. MCI+med. CH n=24, MCI n=11, MCI+med n=15 subjects per group.

### Anthropometric characteristics, physical activity and fitness

All groups were highly active (>10,000 activity counts per day), but differed in physical activity (p=0.010), with MCI+med subjects exhibiting slightly lower activity counts compared to both CH (p=0.008) and MCI subjects (p=0.009). This did not translate into fitness differences; all participants completed a graded cardiorespiratory exercise text (GXT) and overall fitness levels (VO_2 peak_) were not different between groups. However, the duration of the GXT differed significantly (p<0.001) with a shorter test duration observed in MCI+med subjects compared to both CH and MCI subjects (CN vs. MCI, p<0.001, MCI vs. MCI+med, p=0.012). RER was measured in the fasted, rested condition prior to the GXT to assess substrate utilization patterns. Resting RER differed between groups (p=0.031), with lower RER in MCI (p=0.023) and MCI+med subjects (p=0.044), suggesting potential differences in resting substrate metabolism. We also observed differences in fat mass (p=0.027), with MCI individuals exhibiting less fat mass than MCI+med subjects (p=0.007) and marginally lower fat mass compared to CH individuals (p=0.060). Lean mass and bone mineral density were not different amongst groups (**Table 1**).

### Relationship between mitochondrial respiration and fitness

Given the observed group differences in State 3 (ADP) respiration, we examined the linear relationship between State 3 respiration and cardiorespiratory fitness to determine if mitochondrial function is related to cross-sectional cardiorespiratory fitness levels. When we examined only individuals who met American College of Sports Medicine (ACSM) criteria for a reliable maximal exercise test (VO_2max_, n=44 subjects) we observed a positive linear relationship between VO_2max_ and State 3 (ADP) skeletal muscle respiration in both CH subjects (β=0.615, p=0.024) and MCI individuals (β=0.615, p=0.036; **Figure 3**). There was no significant relationship between VO_2max_ and skeletal muscle respiration in medication-treated MCI participants, suggesting a potential dyscoupling of mitochondrial function and fitness in those individuals. We did not observe a significant relationship within the cohort as a whole or within in any diagnosis group when examining the relationship between State 3 (ADP) respiration and VO_2peak_ when including all participants regardless of ACSM criteria, indicating that the exercise test quality is an important factor when investigating these relationships.

**Figure 3.**
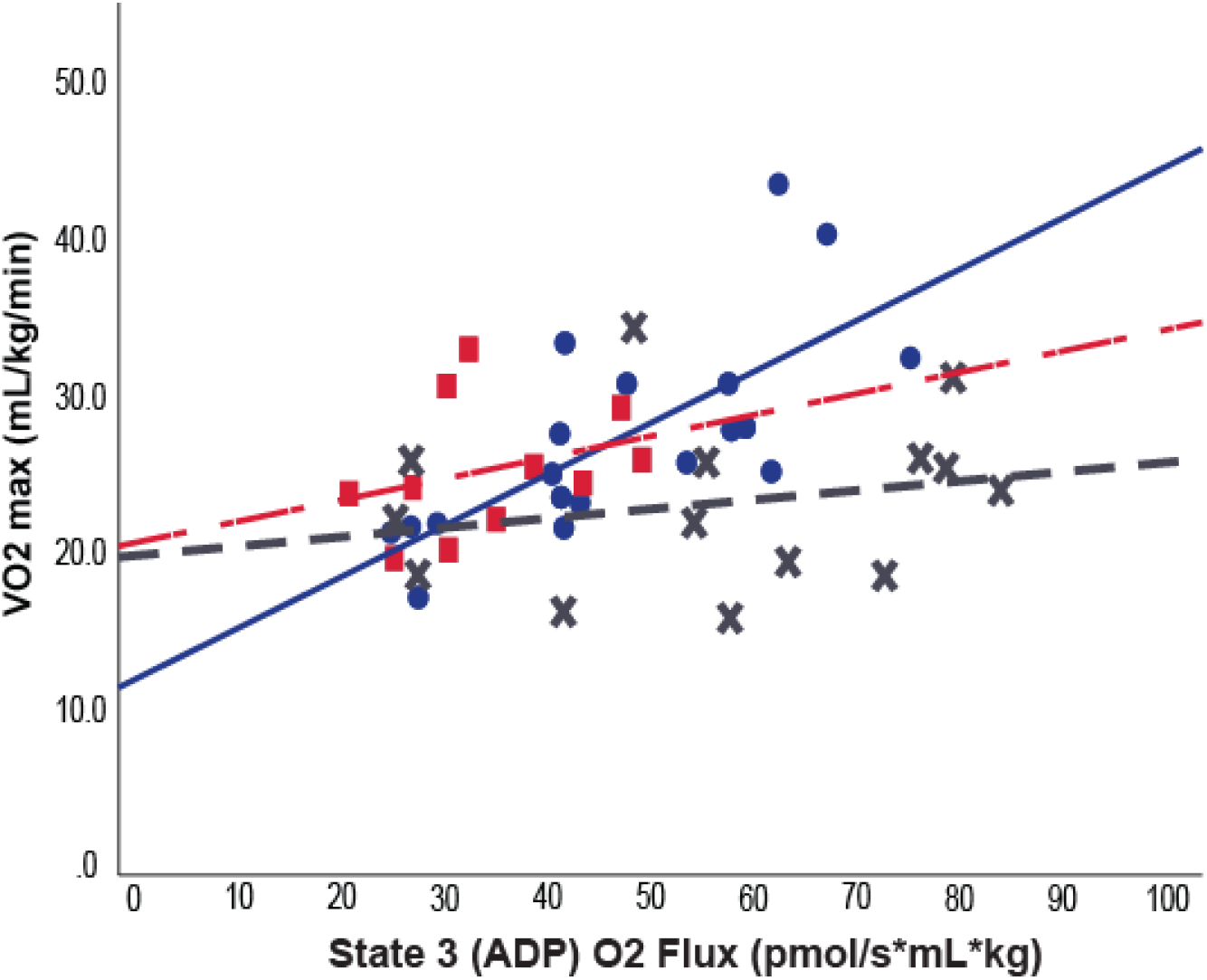
Relationship between skeletal muscle respiration and fitness. In CH (solid blue line) and MCI (dashed red line) individuals who met established criteria for a reliable graded exercise test (n=44), cardiorespiratory fitness tracks with State 3 (ADP) respiration (CH; β=0.615, p=0.024 and MCI; β=0.615, p=0.036). There is no significant relationship in medication-treated MCI subjects (dashed gray line). Relationships were assessed within each group using linear regression, adjusting for covariates. CH n=19, MCI n=11, MCI+med n=14 subjects per group.

### Protein expression of the electron transport system

We assessed protein expression the electron transport system in a subset of participants with sufficient available skeletal muscle samples (n=44). Although complex expression showed the same trends observed for the functional outcomes, these were not significantly different between groups (**Figure 4**).

**Figure 4.**
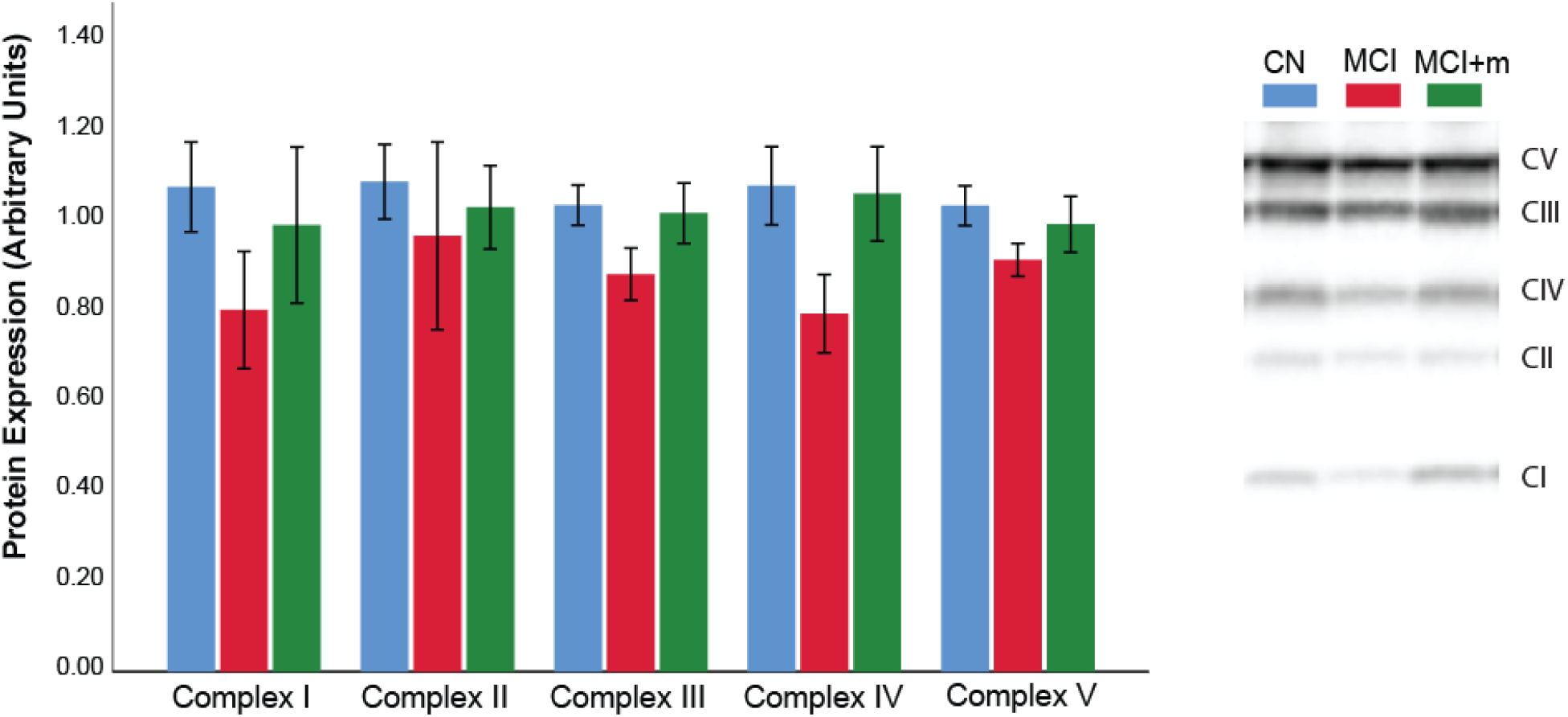
Skeletal muscle protein expression of mitochondrial complexes. Untreated MCI individuals exhibited a consistent but nonsignificant trend for lower protein expression of mitochondrial complex proteins in skeletal muscle. Group differences were assessed using ANCOVA, adjusting for covariates. These analyses were performed on a subset of individuals (n=44) for whom sufficient muscle sample was available (CH n=21, MCI n=10, MCI+med n=14).

### Blood biomarkers

We did not observe group differences for plasma cholesterol, triglycerides, Low Density Lipoprotein (LDL), High Density Lipoprotein (HDL), glucose, or insulin (**Table 2**). LDL/HDL ratio and homeostatic model assessment of insulin resistance (HOMA-IR) did not differ between groups.

**Table 2.**
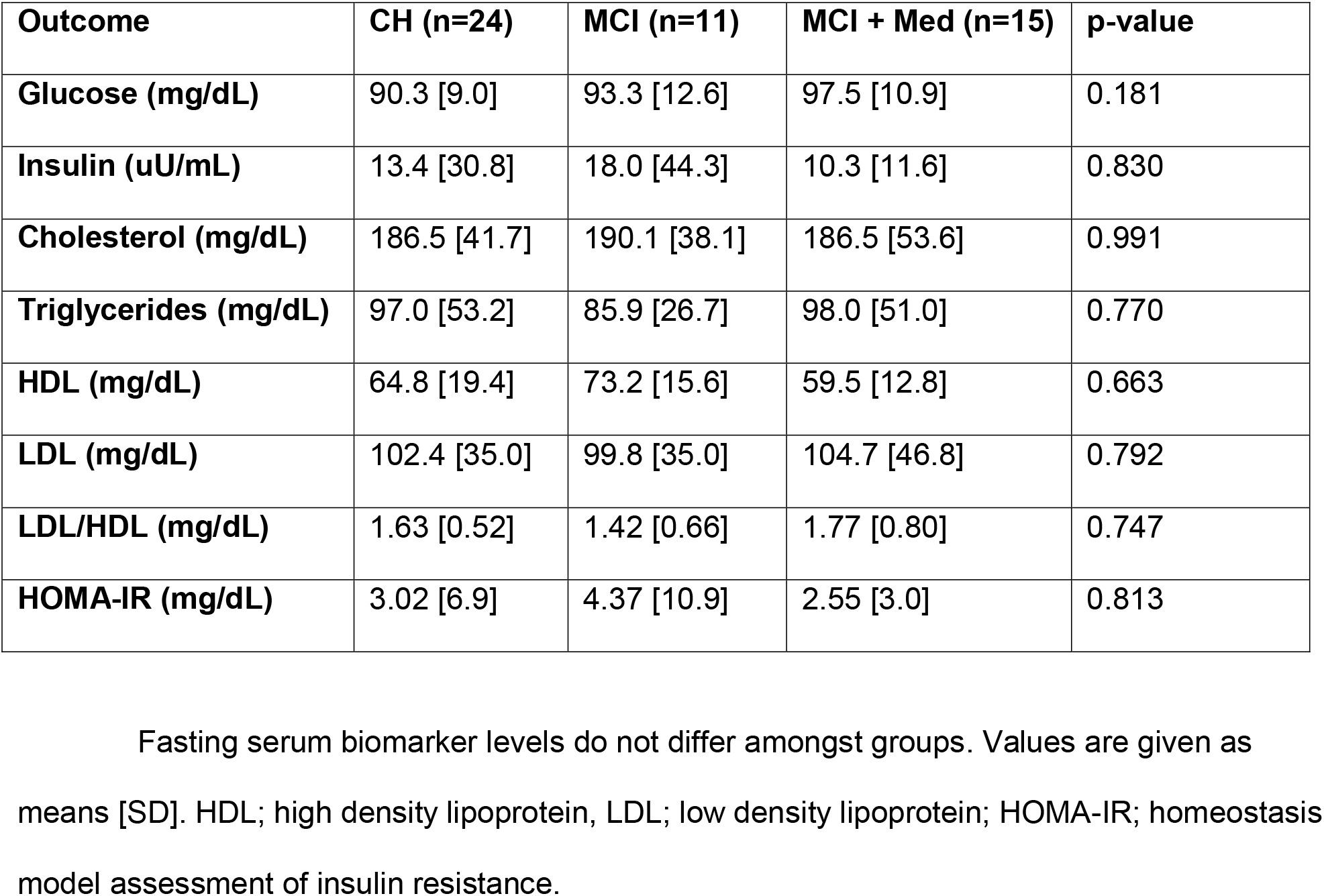
Metabolic biomarkers.

### Adverse events

There were 6 adverse events possibly or definitely related to the GXT or muscle biopsy: 2 mild, and 4 moderate in severity. Mild adverse events possibly or probably related to the intervention included abnormal screening labs (1) and prolonged muscle soreness after the muscle biopsy (1). Moderate severity adverse events included heart rhythm abnormalities during fitness testing (3) and fainting after muscle biopsy (1).

## Discussion

We report here the first ex-vivo assessment of skeletal muscle mitochondrial function in MCI, the earliest clinical stage of AD. Our primary finding is that unmedicated MCI participants display reduced skeletal muscle mitochondrial respiratory capacity (**Figure 1**) and poorer coupling control (**Figure 2**) compared to CH older adults. This suggests that prior findings of reduced insulin sensitivity and cardiorespiratory fitness in AD^2,3^ may be partly driven by deficits in skeletal muscle mitochondrial function. Moreover, MCI subjects treated with AD medication (primarily acetylcholinesterase inhibitors) displayed mitochondrial respiration values in line with CH subjects. Our results thus also suggest that a less-recognized mechanism of medication-related benefit may be improved mitochondrial function in systemic metabolic tissues and the brain. However, future prospective (pre- and post-treatment) studies are necessary to confirm that a commonly used AD medication can indeed impact skeletal muscle mitochondrial dysfunction in MCI.

There is growing evidence for bioenergetic deficits in AD. Cerebral hypometabolism, a marker of reduced energy metabolism, is an early AD biomarker^23,24^ and occurs first in highly metabolic brain regions^25,26^. While bulk of energy in the brain is derived via oxidative phosphorylation in mitochondria, it remains difficult to assess cerebral mitochondrial function directly. However, there is evidence mitochondrial function may be compromised systemically in early AD^1,27,28^. It has also been shown that reactive oxygen species produced in mitochondria enhance Aβ production, which can deposit within mitochondria resulting in further deficits ^28,29^. Preclinical work further suggests mitochondrial deficits in AD models. Mice transgenic for human APOE4 display mitochondrial dysfunction in neurons,^30^ and both the double and triple transgenic AD mouse models^31,32^ display marked mitochondrial dysfunction in skeletal muscle ^32,33^. Our findings of a deficit in mitochondrial respiration in MCI compared to cognitively healthy older adults support a growing body of evidence implicating bioenergetic dysfunction and cognitive decline. To our knowledge this is the first study to directly characterize an ex vivo skeletal muscle mitochondrial function deficit in MCI.

Preclinical evidence also supports our observations of recovered mitochondrial function in medication treated MCI subjects. In our study, all 15 subjects in the MCI+med group were receiving the cholinesterase inhibitor donepezil. In transgenic AD mice, donepezil treatment ameliorates cognitive deficits prior to acetylcholinesterase inhibition and decreases accumulation of Aβ in brain mitochondria.^34^ Pre-treatment of rat brain mitochondria with donepezil also decreases mitochondrial accumulation after oligomeric Aβ exposure and mitigates Aβ-related decline in ATP production, further suggesting protection against Aβ- induced deficits.^34^ Donepezil has also shown anti-amnestic effects and protection against Aβ- induced neurodegeneration when both are administered intracerebroventricularly in mice.^35^ It is possible that some of the benefits of donepezil are mediated through the sigma 1 receptor, a chaperone protein that localizes to mitochondria associated endoplasmic reticulum membranes.^36^ This is of interest because endoplasmic reticulum-mitochondria communication is postulated to be a key factor in AD-related metabolic disturbances and impaired mitochondrial respiration.^37^ Donepezil treatment has further been shown to induce mitochondrial biogenesis in both primary cultures of mouse hippocampal neurons and in hippocampal tissue, an effect that has been linked to induction of AMP-activated protein kinase (AMPK) and peroxisome proliferator-activated receptor gamma coactivator 1-alpha (PGC1α).^38^ Finally, other cholinesterase inhibitors or simply enhancing cholinergic tone has shown direct benefits to mitochondria in cell studies, including improved mitochondrial function, decreased endoplasmic reticulum stress, and prevention of amyloid-beta induced apoptosis.^39,40^

It should be noted that the NMDA receptor antagonist memantine was concomitantly used with donepezil in 5 MCI+med subjects. Memantine has also been shown to stimulate mitochondrial function^41^ and upregulate autophagy, enhancing clearance of damaged mitochondria from cultured neuronal cells.^42^ Thus it is likely that mitochondria-related effects in response to AD medication use may be of clinical relevance. In humans, the symptomatic benefits of donepezil have historically been attributed to effects on acetylcholine neurotransmission,^43^ and it remains debated as to whether cholinesterase inhibitors significantly modify the course of AD. Some degree of neuroprotection has been shown with donepezil,^44,45^ with donepezil treatment associated with slowed hippocampal atrophy.^46^ Moreover, a recent meta-analysis of over 40 randomized clinical trials involving cholinesterase inhibitors found a reduction in mortality, supporting a potential disease-modifying effect.^47^ Positive long term results have led investigators to suggest that cholinesterase inhibitors improve redox balance^48^ and oxidative capacity.^49^ This is supported by the main findings in our study, which show a decline in skeletal muscle mitochondrial respiratory capacity in unmedicated MCI subjects but not in the MCI+med group. The novelty in our findings is that medication treatment is associated with improved mitochondrial outcome measures in skeletal muscle. As already stated, a prospective study with donepezil treatment would be needed to determine if the drug truly rescues mitochondrial function in skeletal muscle. It will also be critical to determine if donepezil is working specifically on skeletal muscle or if is primarily improving neuronal/brain function and changes in muscle mitochondrial function are secondary. Despite the MCI+med group showing improved state 3 respiration, the group still showed reduced coupling and leak control ratio in line with MCI not taking medication. These impaired coupling values are likely due to increased rates of basal proton leak. Future work is needed to examine if altered uncoupling protein 1 or adenonine nucleotide transferase activites are driving increased basal proton leak.^50^

We then examined the relationship between cardiorespiratory fitness and ADP-stimulated mitochondrial respiration in individuals who met established ACSM criteria for a maximal exercise test ^51^ (VO_2max_; n=44). We observed a significant, positive linear relationship between cardiorespiratory fitness and ADP-stimulated mitochondrial respiration in both CH and MCI groups (**Figure 3**). This is consistent with prior work showing that skeletal muscle mitochondrial outcomes (content and function) are associated with whole-body exercise capacity in cognitively healthy individuals throughout the lifespan.^52–54^ In contrast, there was no relationship between cardiorespiratory fitness and mitochondrial respiration within medication treated MCI subjects. This suggests that treatment may dyscouple the mitochondrial function and fitness relationship, although further work is needed to investigate this possibility. Interestingly, mitochondrial respiration was not related to cardiorespiratory fitness in any group when we did not use ACSM criteria for a valid exercise test (VO_2peak_ instead of VO_2max_) – an important consideration for studies examining the relationship between cellular respiration and whole-body fitness.

We anticipated that we would observe differences in skeletal muscle electron transport system protein expression between groups, as prior work suggests cytochrome oxidase deficits in MCI and AD,^28,55^ and a recent meta-analysis has shown complex IV deficits in AD brain.^56^ While we observed trends in the expected direction, these assessments did not reach significance, which could be due to our small sample size in the MCI group (**Figure 4**).

Interestingly, we found that both unmedicated MCI and MCI+med subjects exhibited lower fasting RER values during quiet rest prior to the GXT. Both groups of MCI subjects appear to be more primed to use fatty acid substrates compared to cognitively healthy elderly adults, who exhibit an RER with a greater reliance on carbohydrate use. Somewhat surprisingly, we did not observe any differences in the lipid profile or fasting markers of insulin resistance between groups. This may be due to a variety of factors, including the early disease stage, the high degree of physical activity in all the subjects, the cross-sectional nature of the study, or other factors known to affect lipid homeostasis, such as genetics. Nevertheless, this indicates that the mitochondrial bioenergetic differences can exist outside of or even precede whole-body insulin resistance and/or dyslipidemia.

The primary strengths of this study were the robust ex vivo measures of mitochondrial respiration in skeletal muscle biopsy tissue, which is novel and has never been characterized in humans subjects with MCI. However, muscle biopsies are somewhat invasive and our sample size is limited. This limited our ability to parse out potential interaction relationships between diagnostic, genetic, sex, and anthropometric characteristics, especially in terms of medication use and dose. Future work should further investigate the effects of these important factors. In addition, while we assessed mitochondrial function, we did not measure effects on other more dynamic processes regulating mitochondrial quality control such as mitophagy or mitochondrial fission/fusion. Finally, our study focused on MCI diagnosed in a clinical setting from community-based providers. Additional work on MCI subjects with further characterization, such as harmonized neuroimaging measures, is needed. Nonetheless, our clinically-relevant findings suggest that there is mitochondrial defect in skeletal muscle associated with a disease that occurs in the brain. Furthermore, it suggests a potential new mechanism by which donepezil may impart benefit in early stage cognitive decline.

In conclusion, we show mitochondrial bioenergetic deficits occur in skeletal muscle, a metabolic tissue that is critical for strength, movement, and metabolism particularly in aging, during the earliest stages of AD-related cognitive decline. Moreover, these effects are likely affected by the commonly used AD drug donepezil. Additional work to examine mechanisms by which skeletal muscle mitochondrial dysfunction contributes to AD and how donepezil effects these links is warranted.

## Data Availability

Data available upon request following the publication of this manuscript.

## Author contributions

Study design: J.K.M. and J.P.T.

Conducting experiments: C.S.M., K.N.F., H.M.W., X.W.

Acquiring data: J.K.M., C.S.M., K.N.F., C.S.J., H.M.W., X.W., E.DV.

Analyzing data: J.K.M, E.D.V., P.S., and J.D.M

Manuscript drafting: J.K.M, C.S.M., K.N.F., C.S.J., H.M.W., J.M.B., E.D.V., R.H.S., J.D.M., P.S. and J.P.T.

## Acknowledgements

This project was supported by NIH R21AG056062-01 from the National Institute on Aging. Other support for the authors includes R00AG050490 and R01AG062548 (JKM) R01KD121497 and R01AR071263 (JPT); TL1TR002368 (CSM); K-INBRE P20 GM103418 (JKM, JPT); K99AG056600 (H.M.W), R21AG061548 (EVD), and P30AG035982. We would also like to thank Dr. Chris Perry for technical assistance, and thank our research participants for their invaluable contribution to this project.

## Notes

### Competing Interest Statement

The authors have declared no competing interest.

### Clinical Trial

No trial ID - observational study, not a clinical trial.

### Author Declarations

This study was approved by the University of Kansas Medical Center's Institutional Review Board (IRB # 140787). All participants in this study provided informed consent according to institutional guidelines and in accordance with the Declaration of Helsinki.

